# “We need that consent to protect ourselves!”: An in-depth analysis of health care provider’s perspectives on informed consent and debriefing for caesarean section in West Cameroon using an interpretivist paradigm

**DOI:** 10.64898/2025.12.28.25343105

**Authors:** Jovanny Tsuala Fouogue, Miho Sato, Louise Tina Day, Mitsuaki Matsui, William Carter Djuatio Kenne, Bruno kenfack, Lenka Beňová, Veronique Filippi

**Affiliations:** Department of Infectious Disease Epidemiology and International Health. London School of Hygiene & Tropical Medicine. Keppel Street, London WC1E7ht, United Kingdom; School of Tropical Medicine and Global Health. Nagasaki University. 1-Chome_12-4 Sakamoto, Nagasaki,852-8102, Japan; Department of Public Health, Kobe University Graduate school of Health Sciences. 1-1 Rokkodai-cho, Nada-Ku, Kobe 657-8501 Kobe, Japan; Regional Ethics committee for Human Health Research. PO Box: 479, Bafoussam., Cameroon; Department of Obstetrics, Gynecology and Maternal Health. Faculty of Medicine and Pharmaceutical Sciences, University of Dschang. PO box: 96 Foto Hill, Dschang Cameroon; Department of Public Health, Institute of Tropical Medicine. Nationalestraat, 155, Antwerp. Belgium; Department of infectious Disease Epidemiology and International Health. London School of Hygiene & Tropical Medicine. Keppel Street, London WC1E7ht, United Kingdom

**Author notes:** **Corresponding author full address.** Jovanny Tsuala Fouogue, P.O. Box: 960. Kamkop (Entrée Cooperant), Bafoussam. West-Cameroon., **Corresponding author email address.** Jovanny Tsuala Fouogue.

**Keywords:** Perspectives, Healthcare providers, Informed consent, Postoperative debriefing, Cameroon, Caesarean section, Quality

## Abstract

**Introduction:** Women-centredness is a key quality domain for caesarean section (CS) which can be strengthened by adequate informed consent and post-operative debriefing. CS providers are instrumental in delivering these services and in appraising their adequacy, along with facilitating factors and barriers. However, little is known about providers’ perspectives on informed consent and debriefing practice in countries with limited access to and quality of CS, like Cameroon.

**Objective:** To explore CS providers perspectives on the routine practice of informed consent and post-operative debriefing in West Cameroon.

**Methods:** From March to August 2024, we conducted face-to-face in-depth interviews with 69 CS providers purposively selected to reflect all clinical specialties, gender, levels of seniority and sociolinguistic profiles. Participants worked in the twenty hospitals in the West Region of Cameroon that recorded at least 100 CSs in 2022. Using an interpretivism paradigm, we generated codes inductively from verbatim transcripts and conducted thematic analysis.

**Results:** Three themes emerged: Domination of professional motivation for consent and debriefing by fear of litigations and the need to address unhealthful behaviours at the expenses of genuine women-centredness; Widespread multifaceted structural, processual and staff-related shortcomings in hospitals thwarting professional opportunities to deliver women-centred informed consent and debriefing; The negative impact of socio-cultural reluctance and misinformation on the provision consent and debriefing.

**Conclusion:** Building on the multidimensional shortcomings reported by CS providers, a quality-improvement intervention towards women-centredness of informed consent and debriefing for CS could be designed targeting professional behavioural changes through standardized practice guidelines and supportive oversight among others.

**Highlights:**

- We report care providers’ views on consent and debriefing for c-section in Cameroon
- Health provider’s major motivation for consent and debriefing was fear of lawsuits
- Health provider’s minor motivation was the need to address unhealthful behaviours
- Health providers found hospitals’ context unconducive to consent and debriefing
- Social norms and misinformation hamper proper consent and debriefing for c-section

## 1. Introduction

Bodily autonomy in decision-making regarding reproductive health lies at the core of women’s sexual and reproductive rights and obliges healthcare providers to seek informed consent (IC) prior to every procedure (1,2). Under relevant legal frameworks, international and national public health and professional bodies have established comprehensive regulations and guidelines governing the practice of informed consent for various reproductive health procedures (3–7). The foundations of these guidelines, as grouped by Beauchamp and Childress (2013), fall into three categories: women’s competence and capacity, adequate disclosure and understanding of relevant information and authorization (8). However, from the perspective of women’s need for communication about what has happened to their bodies, focusing on informed consent might overshadow the need for postoperative debriefing (D). Such debriefing is essential to complete the provision of information on caesarean section (CS) (9–13)

CS distinguishes itself from other reproductive health procedures, due to its high frequency among major surgeries worldwide, its powerful symbolic association with procreation, and its key contribution to the reduction of maternal and newborn morbidity and mortality, particularly in low- and middle-income countries (LMICs) (14,15). While improving access to emergency obstetric care remains an important programmatic area in these countries, there is a growing focus on improving the quality of obstetric care, so that it becomes more women centred. This includes ensuring informed and consented care for CS within health-service strengthening agendas for mothers and newborn(16). However, in sub-Saharan Africa (SSA) the evidence-base remains limited for informed consent for CS and virtually inexistent for debriefing (12)

Maternity care providers operating at the interface between health systems and women play a critical role as street-level bureaucrats in the delivery (or lack thereof) of policies on informed consent and post-operative debriefing for CS (17). Their proficiency must be adequate, and their attitudes supportive, to ensure the effective enactment of governmental policies on those issues. Additionally, their perspectives are essential for understanding the factors that enhance or undermine the effectiveness and quality of these services.

Nevertheless, we identified only nine articles describing the perspectives and attitudes of Africa-based healthcare providers from various clinical backgrounds on informed consent for surgery in general in Nigeria, Somaliland, Tanzania, South Africa, Malawi and Uganda (17–25).

Cameroon is a lower-middle-income SSA country with a total fertility rate of 4.8 births per woman and a CS proportion of 3.5% in 2018 (26). In 2022, the Ministry of Health enacted the current version of the national code of medical ethics, which introduced the compulsory requirement of informed consent for every medical procedure or examination (27). Regarding CS, the only published research on informed consent in the country revealed that only half of the women who signed a consent form for CS at a national reference hospital between 2020 and 2021 did so after receiving verbal information (28). In 2023, we conducted an audit of the documentation of informed consent and debriefing in the 23 hospitals that performed two-thirds of all CSs in the West Region of the country. In our sample of 234 CSs only 36.8% of informed consent and 0.4% of debriefing cases were documented (Fouogue et al. 2023. Unpublished results).

Given the limited data on informed consent in Cameroon and other African settings and evidence that it is poorly practiced, we conducted a qualitative study to gain an in-depth understanding of CS providers’ perspectives on informed consent and post-operative debriefing.

## 2. Methods

### 2.1. Researchers’ characteristics and reflexivity

#### 2.1.1. Personal characteristics and relationship with participants

A lead researcher (JTF) and a research assistant (WCDK) collected and analysed the data. JTF is an obstetrician gynaecologist and public health physician; he is a university lecturer and senior consultant at the reference hospital in the West Region of Cameroon. At the time of the research, he was enrolled in a joint PhD programme at the University of London and Nagasaki University. WDCK is a social health scientist who previously worked as a consultant for several development non-governmental organizations in Cameroon. WCDK researchers had no previous interactions with participants; Likewise, JTF had no previous interactions with the included women but had collaborated professionally with approximately one-sixth of the included healthcare providers. The latter were aware of his role in conducting research to improve the quality of health services.

#### 2.1.2 Reflexivity statement

Lead researcher (JTF): « I am a Black, able-bodied Cameroonian man with a Bamileke cultural heritage. My socio-professional privileges stem primarily from my higher education and gender. Reflecting on my 15-year experience conducting CSs in four regions of Cameroon and Switzerland, I recall witnessing, contributing to, or mitigating deplorable CS consent procedures. These practices affect women across all socioeconomic categories. I believe that CS providers too often use resource limitations as a convenient excuse to justify their inaction in fostering women’s autonomy and improving the quality of care. Women too often receive disrespectful, non-compassionate, and undignified care. Healthcare providers frequently adopt a paternalistic approach toward female patients, which is deeply troubling. Rooted in progressive political values, I believe this pattern is largely driven by a lack of downward accountability, a system in which those in positions of power are not answerable to the users of their services. Just as some civil servants may not feel accountable to the public, healthcare providers often do not feel directly accountable to their patients. This is particularly evident in interactions with women, who commonly have deferential attitudes towards healthcare providers.

I recognize that my personal convictions, male background and experience may influence the way I interpret the data, by amplifying barriers to women-centeredness. Furthermore, my current position as a public health physician in the West Region of Cameroon could have triggered a socio-professional desirability bias during interviews, particular with participants affiliated with the health system. »

### 2.2. Context and study sites

#### 2.2.1 Context

The West Region of Cameroon is one of the ten administrative regions (29,30). It is divided into 20 health districts, and the population was 2,398,224 in 2024 (31). In 2018, almost all deliveries (96.9%) were facility-based. The regional prevalence of CS was approximately 10% in 2023, the procedure was performed in 96 hospitals, each conducting between 1 and 500 CSs per year (32). About 80% of CSs were emergency procedures in 2022 (33). Household direct out-of-pocket payment CS remains prohibitive and caused catastrophic health expenditures in 42% of cases in two neighbouring regions in 2023 (34). Despite a high regional literacy rate, the gender inequality index is high, and patriarchy is deeply rooted in the collectivist social fabric of communities (35,36). The region is home to semi-Bantu ethnic groups, the largest being the Bamileke followed by the Bamoun; Ethnic minorities include the Fulbe, Tikar, and Mbo. Ethnic groups are organized into 127 strong traditional kingdoms. These kingdoms are sociopolitical and geographic entities centred on kings who are paramount political and spiritual leaders believed to wield divine power. Regarding official religions, Bamileke and Mbo are predominantly Christians, while Bamoun, Tikar, and Fulbe are mainly Muslims (37). Bamileke and Bamoun traditional religions which mainly consists of the ancestors’ worship and several moral bans, remain widely practiced alongside modern religions.

#### 2.2.2 Sites

We systematically included hospitals in the West Region of Cameroon that recorded 100 or more CSs in 2022. Of the 20 hospitals that spanned 9 of 20 health districts of the Region, three were rural and 14 were public while six where Christian faith based private. Regarding their levels, three were sub-district hospitals, 14 were district hospitals, and three were regional-level hospitals. The resulting distribution encompassed all the ethno-geographic components of the Region.

### 2.3 Study design

#### 2.3.1 Qualitative approach, guiding theory and research paradigm

We conducted a thematic analysis of CS providers’ perspectives, obtained during IDIs. We interviewed CS providers individually because they formed a diverse group of specialized professionals (nurse assistants, nurses, midwives, surgeons, obstetricians, scrub nurses, and anaesthetists) working in several units and departments, often within complex multidisciplinary and hierarchical structures. We deemed IDIs appropriate for in-depth exploration of perspectives within each speciality and to minimise the influence of professional power dynamics on participants’ openness. Moreover, conflicts between specialities and units that potentially affect informed consent and debriefing were expected to be freely recounted.

We adopted an interpretivist research paradigm. Interpretivism upholds that truth and knowledge are subjective as well as historically and culturally situated, based on people’s experiences and their understanding of them (38). It emphasises individual agency and interpretation within social structures and institutions. This aligns with the goal of this study.

#### 2.3.2 Participant selection

We conducted purposive sampling of healthcare providers directly involved in the CS continuum of care, from antenatal clinics to postnatal consultations. We aimed to recruit enough participants to attain data saturation and exhaustive coverage of the 20 selected hospitals (39,40).

#### 2.3.3 Data collection, processing and analysis

We designed and pretested English and French versions of the guides used to conduct interviews between March 24, 2024, and August 29, 2024. We audio-recorded the interviews with the participants’ permission. In addition, we systematically took field notes during the interviews. JTF and WCDK checked the quality of the audio recordings and anonymised them before proceeding with transcription as and when interviews were carried out. We also verified the initial transcripts for accuracy and revised them when necessary.

Using NVivo® 14 software (Lumivero, LLC), we (JTF and WCDK) inductively developed codes. After coding the first four transcripts, we elaborated a joint codebook. To enhance the trustworthiness of the data, we performed constant memoing, systematic reference to field notes, and cross-comparisons between coders. Saturation was reached with respect to the proceedings (processes, actors, materials and content) of consent and debriefing after coding the transcripts from 12 of the 20 hospitals. However, new meanings related to contextual sociocultural features kept emerging, leading us to continue data collection and coding for all the remaining hospitals (39,40).

JTF edited the harmonised codebook that was further discussed by the research team (VF, MS, LTD and MM). Building on the final codebook, we generated initial themes through an analysis of emerging patterns and the identification of relationships namely contradiction, similarity, causality, hierarchy, co-occurrence, and intersection. This initial list of themes was reviewed and reframed together with the research team (VF, MS, LTD and MM). In this paper, the results are presented in accordance with two guidelines: the Standard for Reporting Qualitative Research and the COnsolidated guidelines for REporting Qualitative research (COREQ) (41,42).

### 2.4 Ethics

This study was approved by the Research Governance and Integrity Office of the London School of Hygiene and Tropical Medicine (LSHTM Ethics Reference: 29898) and the Regional Ethics Committee for Human Health Research for the West Region of Cameroon (Reference: N°/984/25/10/2023/CE/CRERSH-OU/VP). We observed all the requirements of the World Medical Association’s Helsinki Declaration (version 2013) on research involving humans and Cameroon’s relevant legal requirements. The transcripts were anonymised and all participants’ identifying information were removed.

## 3. Results

Figure 1 depicts the inclusion criteria for the participants. All 69 CS providers approached consented to participate.

**Figure 1.**
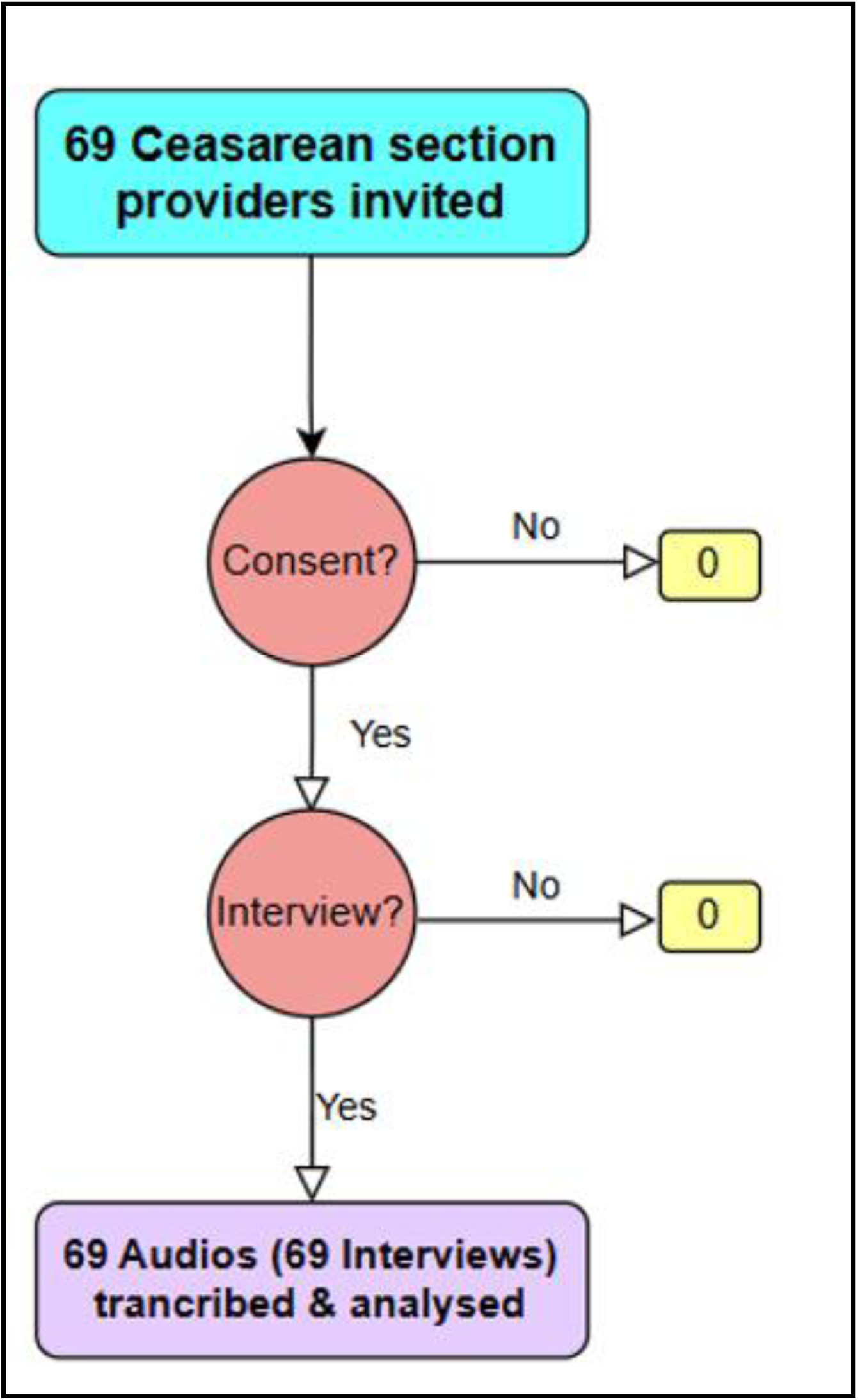
Flow diagram: inclusion of the participants. Health care providers’ perspectives on informed consent before caesarean section and debriefing thereafter. West Region of Cameroon. N = 69. March – August 2024.

### 3.1 Sample characteristics

Table 1 summarizes the participants’ characteristics of. The median age (range) was 38 (23–59) years, and the median duration (range) of the IDIs was 33.5 (14.15 – 89.45) minutes. Half of the participants were women, and approximately two-thirds of them worked in maternity units.

**Table 1.**
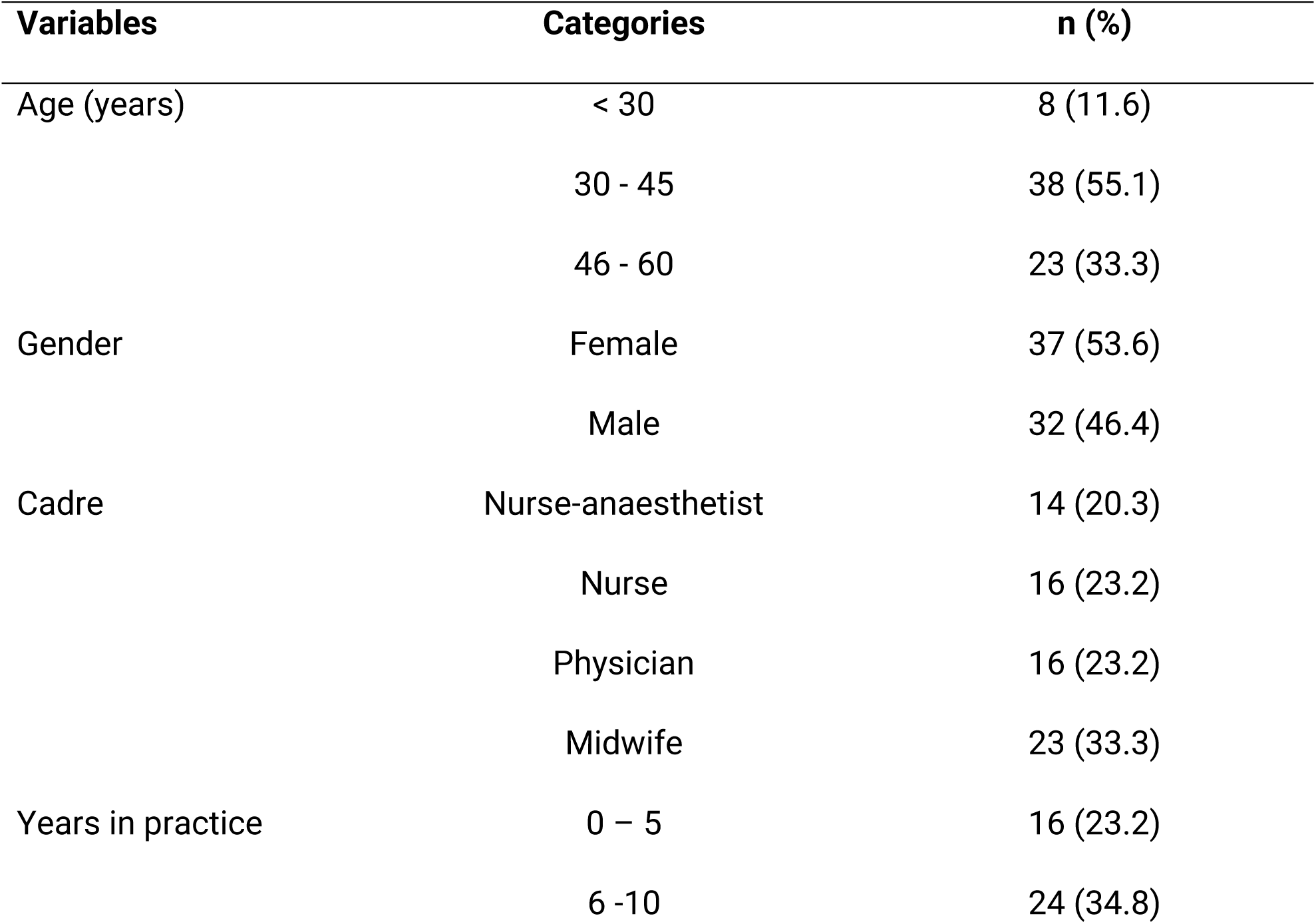
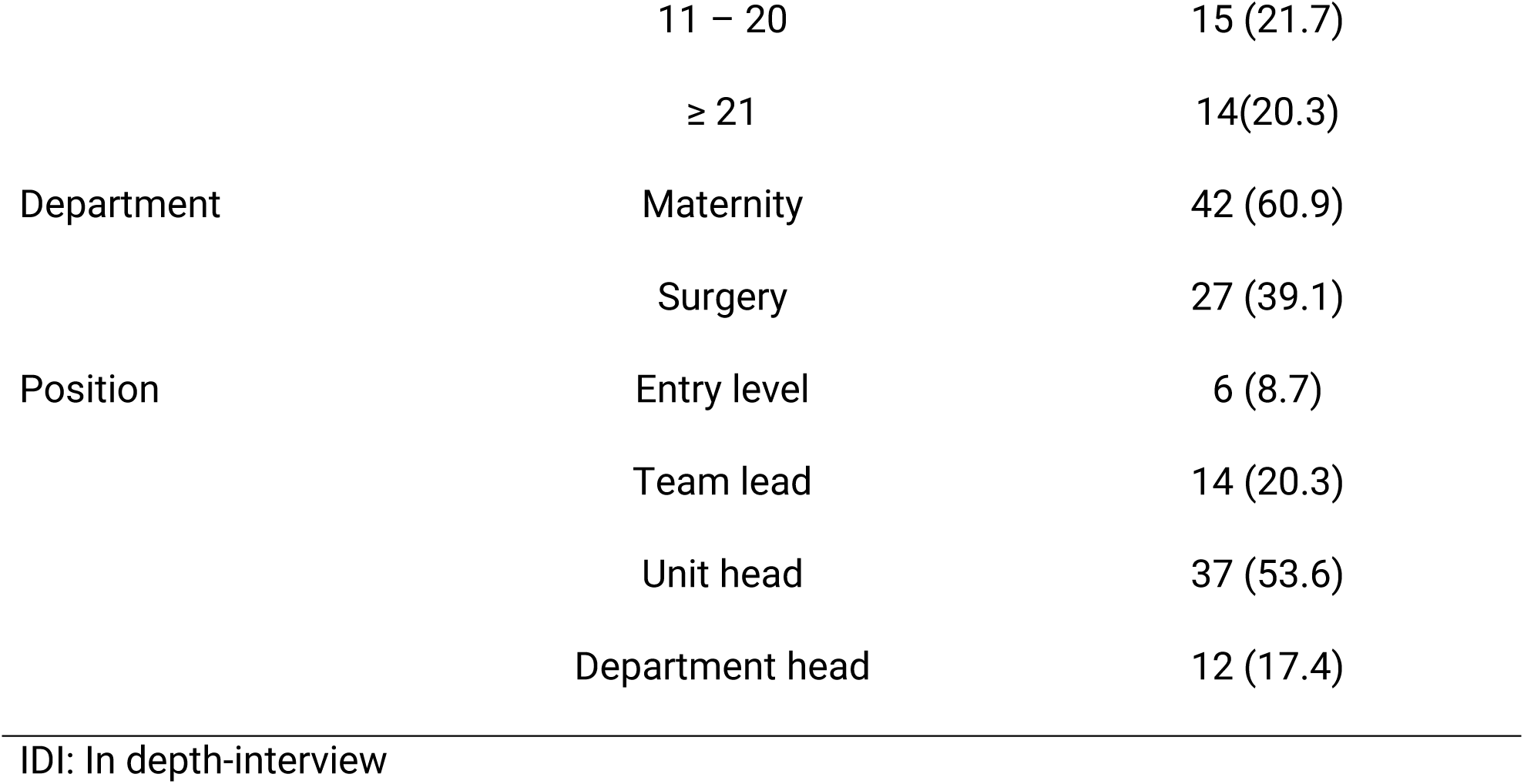
Sample characteristics. Healthcare providers’ perspectives on informed consent and debriefing after caesarean section. West Region of Cameroon. N=69. March – August 2024.

| Variables | Categories | n (%) |
| --- | --- | --- |
| Age (years) | < 30 | 8 (11.6) |
|  | 30 - 45 | 38 (55.1) |
|  | 46 - 60 | 23 (33.3) |
| Gender | Female | 37 (53.6) |
|  | Male | 32 (46.4) |
| Cadre | Nurse-anaesthetist | 14 (20.3) |
|  | Nurse | 16 (23.2) |
|  | Physician | 16 (23.2) |
|  | Midwife | 23 (33.3) |
| Years in practice | 0 – 5 | 16 (23.2) |
|  | 6 -10 | 24 (34.8) |
|  | 11 – 20 | 15 (21.7) |
|  | ≥ 21 | 14(20.3) |
| Department | Maternity | 42 (60.9) |
|  | Surgery | 27 (39.1) |
| Position | Entry level | 6 (8.7) |
|  | Team lead | 14 (20.3) |
|  | Unit head | 37 (53.6) |
|  | Department head | 12 (17.4) |

IDI: In depth-interview

### 3.2 Themes

From the 69 transcripts, we generated three themes:

- Motivation to provide informed consent and debriefing dominated by the mere fulfilment of legal requirements and the need to address unhealthful CS-related practices.
- Acknowledgement of widespread inadequate informed consent and debriefing practices grounded in unconducive hospitals environments.
- Negative impact of socio-cultural hostility, low educational attainment, and misinformation on the provision informed consent and debriefing.

#### 3.2.1 CS providers’ motivation to provide informed consent and debriefing

CS providers mainly perceived provision of informed consent and debriefing as the mere fulfilment of legal requirements and an opportunity to address misconceptions and harmful practices related to CS.

##### 3.2.1.1 Fulfilment of a legal requirement

CS providers presented informed consent and debriefing as instruments to ensure women’s right to receive relevant information mostly as passive recipients before and after an invasive procedure, such as CS, and exercise their right to refuse. They situated the right to informed consent and debriefing within the provision of dignified care. Beyond this, they stated that informed consent and debriefing ensured their protection against eventual postoperative violence from family members, outrage on social media and even malpractices litigations that are on the rise in the country (43–46). When a complication or even death occurs, which is more frequent with delayed referrals preventing informed consent to be sought in receiving hospitals, family members typically blame the referral hospital instead of the whole chain of (often unconventional or unlicensed) providers upstream. They further explained that while informed consent could be waived or expedited for several reasons (incapacity caused by critical pregnancy and labour complications and extreme life-threatening emergencies, consented planned CS ultimately done in emergency), there was no justification to discharge a woman home without appropriate debriefing given the crucial care and support women and babies needs after a CS. These included counselling on physical, dietary, and reproductive restrictions required during the recovery period, alongside education on danger signs and general postnatal advice. They made a special case for the strict observance of the interpregnancy interval, which requires the use of family planning methods and husbands’ cooperation.

> *“I tell them that there could be consequences, and we find ourselves in court. Moreover, the world had evolved, and we would like to have your opinion; I can no longer operate you like if there were animals that one takes, puts on a table and does; No, we must agree before.” An **anaesthetist nurse***
>
> *“Just imagine a woman who goes somewhere [low level or unconventional health facility] with a scar uterus and gets killed [uterine rupture] because she did not observe the recommend interpregnancy interval or had a contra-indicated labour. If they [auditors] trace back here and discover she was never informed on anything! Yes, we will be held liable. We need that consent to protect ourselves!” **An anaesthetist nurse***.

##### 3.2.1.2 Opportunity to address socio-cultural practices detrimental to CS

CS providers presented informed consent and debriefing as opportunities to address widespread misbeliefs and misinformation about CS because women and families are keener to listen at that time. For planned CS, midwives led the antenatal consultations. They insisted on the constant need to involve husbands and women’s in-laws and listed several key topics for discussion: avoiding attempts at high-risk medically contraindicated vaginal births encouraged by non-conventional healthcare providers (traditional birth attendants, herbalists, faith healers); putting money aside to afford an eventual CS, given the out-of-pocket funding model; addressing the stigma that women may face after a CS; reducing excessive fear of deadly complications of surgery and anaesthesia, and building trust in the hospital team by emphasising truthfulness of CS indications in referral hospitals, despite their poor reputation due to their high CS rates perceived as driven by financial reasons. For emergency CS, the previous aspects work with late referrals of women to make informed consent more sensitive, thereby requiring culture-sensitive communication skills and strong work ethics while acting swiftly to save the woman and her baby.

> *“The cultural context is a heavyweight … They first consult a fortune teller! When they arrive at the hospital, in the operating theatre we see women with recent scarifications and ointments made because they were told there should be the informed consent [approval] of the witch doctor before the woman goes to the hospital. Women are still very attached to their culture, their customs, and their traditions. They believe that we operate just for the sake of operating.” **An anaesthetist nurse***.

Debriefing was seen as adequate for counselling on post-natal cultural practices detrimental to post-CS healing. They presented it as critical given the high prevalence of unhealthy post-CS practices. The main reasons included precocious conception, wound washing using herbs, precocious resumption of heavy physical activities, and unclear timeframe for resumption of sex. CS providers insisted that it was paramount to ensure the adoption of a contraceptive method to observe the recommended post-CS interpregnancy interval.

> *“While discharging them home, we explain the relevance of the CS they had and what lies ahead if they don’t observe the recommended interpregnancy interval.” **A scrub nurse***.

#### 3.2.2 Acknowledgement of widespread inadequate informed consent and debriefing practices

CS providers deplored several shortcomings in the practices of consent and debriefing that we described elsewhere (Fouogue et al. 2025. Unpublished results). Consent for elective CS is sought during antenatal consultations by the attending midwife from the woman who always secures her husband’s approval before notifying her final decision. In emergency CS, providers mainly engage with accompanying relatives who in turn refer to husbands, sometimes completely bypassing the woman herself. Debriefing after CS is not done in public hospitals and is content-limited in private faith-based hosptials. We grouped their criticisms into three categories: hospital structures and processes, providers’ background and contents of consent and debriefing interactions.

##### 3.2.2.1 Hospital structural inadequacies

Providers cited a lack of physical spaces for confidentiality and regulatory instruments (standardised procedures, forms, monitoring). In addition, the rare consent forms (mainly in faith-based hospitals) and the sole debriefing form were criticized by CS providers for poor content both technically and legally. CS providers in most facilities deplored that debriefing was ineffective and that they failed to give post-CS women the recommended procedure-specific information.

> *“During rounds we do it superficially, but there’s no structured framework, and it’s not really standardized. I think that’s a weakness we need to work on”. **A general practitioner***

##### 3.2.2.2 Process inadequacies

In addition, to their non-standardized nature, informed consent processes were widely described as being based on an opt-out approach: women and their accompaniers either consented to the CS and stayed in hospitals or refused and had to leave. Providers outlined their hurry to refer or drive away reticent women, given that most CS are emergency procedures that are prone to complications, scandals, and litigations. Before leaving the hospital, the women and their accompaniers were compelled to sign a refusal form. This trend of hasty referral was more marked in large referral hospitals owing to the higher proportion of delayed emergency referrals of women in critical condition from lower-level health facilities.

> *“The woman tells you no, she doesn’t want and will go to another institution. Because in our institution, the patient is not a prisoner; you can come in and leave as you wish. So, if you refuse to give informed consent, you can sign off the discharge against medical advice*.
>
> *You’re free to go wherever you think they’ll agree to operate on you without informed consent.” **A midwife***

According to CS care providers, informed consent transactions would have been smoother if antenatal care providers consistently informed women that CS remains plausible (even) when the vaginal route is the anticipated mode of childbirth. They argued that it would have lessened women’s reticence and suspicion regarding financially motivated CS indications. Finally, in some remote settings, women’s and their accompaniers’ low educational attainment was a communication barrier, necessitating additional human resources for interpretation.

> *“Generally, it’s difficult. There should always be an interpreter. That’s the case for most women who haven’t been to school… it also makes the work a bit more burdensome.” **An anaesthetist nurse***

##### 3.2.2.3 Actors’ inadequacies

CS providers described staff as overworked, job-dissatisfied, and unskilled to adequately handle informed consent and debriefing. They insisted that both informed consent and debriefing are time-consuming and thus human resource-intensive activities that were not prioritized in task distribution by hospital managers. In addition, CS providers said they lacked skills specific to consent and debriefing, namely psycho-emotional support and communication techniques. Only CS providers from public hospitals blamed job dissatisfaction; for them, painstaking, patience-demanding, and compassionate tasks, such as seeking informed consent and providing debriefing, are not properly carried out because providers on site are commonly bitter and frustrated at their working conditions.

> *“We need to look at the things that hinder staff commitment to consent and debriefing. There’s the workload, the patience, and salary-related demotivation!” A **general practitioner***

CS providers also reported inadequate staff allocation. Providers in labour and emergency units responsible for consent in emergency scenarios deplored that staffers doing the CS (obstetricians, surgeons, anaesthetists, and medical officers) were rarely involved in consent discussions because they only arrived once women were in the operating room. To them, this unfairly waived their liability. Likewise, maternity staff in some hospitals were frustrated by being excluded from post-CS care, which was exclusively done by surgery department staff. They felt that debriefing in those cases was therefore suboptimal and that the continuum of maternal care was interrupted.

> ***“****We are not authorized to do it. Yes, it is the gynaecologist himself who must obtain the consent. But when there is an emergency and the gynaecologist is not available, we are forced to do it.” A **midwife***

Providers perceived third-party additional consent as a strongpoint because it aligns with socio-cultural norms, which in turn fosters acceptability and enhances out-of-pocket payments. However, this also shifts decision-making away from women themselves.

> *“For that emphasis on the husband? There is the cultural aspect. In this community, which is predominantly Muslim, the man is the Lord of the household to summarize. Everything goes through him, coming in or going out. So, he truly needs to be aware, informed, and convinced if you want things to work. … If you convince the man without convincing the woman, you’ve convinced both. But if you convince the woman without convincing the man, you haven’t even done 20% of the work. So, the man must be informed—or at least his father. A male person in the community.” **A general practitioner***

##### 3.2.2.4 Content inadequacies

Both informed consent and debriefing were described as quite limited in content in most hospitals. Specifically, providers justified not delving into the risks of potential complications while seeking consent because that would undoubtedly further discourage already fearful and reluctant women and accompanying relatives who never want to tempt fate by mentioning a CS. In addition, the anaesthetists acknowledged that women were rarely given a choice over the technique because loco-regional anaesthesia demands resources that are not available around the clock in most hospitals. During debriefing, which CS providers also referred to as post-CS counselling, obvious surgical complications under management were rarely acknowledged as such to avoid blame and litigation. CS providers further noted that family planning, to help ensure a safe minimum interpregnancy interval, was not systematically offered in many settings.

> *“When she asks questions ‘How is it going to happen? I’m scared’—then we explain. But we don’t talk about the risks; we focus instead on the benefits.” **A midwife***.

#### 3.2.3 Negative impact of socio-cultural hostility, low educational attainment, and misinformation on the provision informed consent and debriefing

Providers revealed that the default response of communities towards CS was either a firm refusal or strong reluctance which complicated the processes of obtaining informed consent and often led to protracted deliberations among women and their support networks, particularly husbands. They also described frequent disruptions of consent discussions as hesitant accompanying relatives sought alternative interventions from non-conventional health providers (traditional healers, faith-based healers, herbalists, and witch doctors) to overturn the indications for emergency CSs and permit vaginal birth, sometimes even within the labour room.

> *“Generally, it’s extremely frustrating, because we have cases where there’s a real emergency, and what the husband decides to do is go look for a traditional healer. I’m talking, for example, about cases of obstructed labour, and the husband insists that the delivery must happen vaginally. Or when there’s a malpresentation, and they tell you a traditional healer will give some medicine that will turn the baby inside the womb. We see cases like this very frequently. He doesn’t see the importance of intervening at that moment, and that leads us straight into complications because if he hasn’t given his decision, you can’t act, you can’t do anything. So, all we can do is insist and try to make him understand the importance of the procedure, which is difficult.” **A midwife***

Many CS providers insisted that one impact of baseline hostility and stigmatisation of CS in communities is the need for psychological and emotional support before and after the procedure. Considering the scarcity of clinical psychologists in hospitals, they suggested the recruitment and involvement of those specialists in the continuum of CS from consent to debriefing.

> *“What I would suggest is having psychological services to support us sometimes. Because if there were people to prepare these women before they’re handed over to us, it would make things easier. And if there are no trained psychologists, then we could train some of the staff on the psychology of pregnant women.” **A midwife***

Supplement 1 presents additional relevant quotes pertaining to all above themes.

## 4. Discussion

We explored the perspectives of CS providers in the West Region of Cameroon on current practices of informed consent and debriefing. Findings revealed that CS providers engaged with informed consent and debriefing not to centre CS care on women but to protect themselves against litigations and counsel communities; Their reports revealed how hospital are structurally unconducive to women-centred informed consent and debriefing; Finally, the hostility of socio-cultural norms towards CS negatively impacted informed consent and debriefing services.

### 4.1 Results in context

#### 4.1.1 Professional motivation and opportunity for provision of informed consent and debriefing

Most CS providers were vocal about the medico-legal function of informed consent and debriefing. This aligns with results from three studies in neighbouring Nigeria, which found that two-thirds of providers were also aware that informed consent protects them from litigation (17,21,22). A possible reason of CS provider’s over-hope on signed informed consent and debriefing to protect them is the recent series of scandals and lawsuits against CS providers in Cameroon (47–50). These high-profile controversies related to CS consent and maternity care led to the imprisonment of several health care providers. CS providers could have therefore adopted a more cautious and defensive approach in their daily practice to adjust to these societal changes. The routine practice of hastily referring or driving away women who refuse emergency CS (“providers’ way or the highway”) illustrates their heightened cautiousness. The unethical nature of this practice must be emphasised. Both Cameroon’s and the World Medical Association’s codes of medical ethics state that providers should continue to attend to patients who refuse medically indicated procedures after ensuring that they have fully understood the consequences of their refusal (27,51). Volatile malpractice environments don’t necessarily correlate with high quality of informed consent and debriefing but are causative of defensive clinical practices among high-liability medical specialties like obstetrics/gynaecology (52). Though the prevalence of malpractice lawsuits remains relatively low in Cameroon the magnifying effect of mass and social media could have impacted CS providers’ behaviour (53) .

There is a concerning mismatch between providers’ motivation to offer informed consent and debriefing and women’s mitigated experience of CS. Indeed, addressing the socio-cultural barriers to CS and avoiding lawsuits are sensible motivations for providing informed consent and debriefing but, more is expected from CS providers to deliver dignified and genuinely women-centred experience of CS care which is a unique combination of high stakes and meanings for women: birthing experience, surgical and general anaesthetic risks, and most often in West Cameroon, traumatic emergencies including referral trajectories/pathways and stigmatisation. Health care providers are called to embody provision of positive experience of CS care beyond merely “wanting” to deliver on that women’s right; they are expected to “need” it to fulfil their duty (54). Indeed, merely “wanting” stems from reflective professional motivation and would only lead to “mechanistic” technically sound delivery of informed consent and debriefing. On the other hand, “needing” it is a product of automatic motivation and will ensure compassionate women-centred services (54).

With few exceptions, CS providers from all our study hospitals provided details on major inadequacies in their routine provision of informed consent and debriefing. Similar quality gaps have been reported in several sub-Saharan African countries. In a South African study, health care providers cited excessive workload, language differences, and time constraints as barriers to adequate informed consent (24). They demonstrated that informed consent was of higher quality when offered by specialist doctors than by other providers (24). A study in Uganda reported a similar routine of informed consent being sought by providers other than those who performed the surgery or being signed after the surgery (23). Shortages in health workforce pose the additional dual challenge of task shifting and competence of non-physician providers to offer informed consent (55).

CS providers revealed that post-operative debriefing was even rarer than informed consent in their respective hospitals. This is a serious shortcoming because adequate delivery of postoperative communication to post-CS women during their hospital stay would solve two issues: customizing post-natal care to CS by including procedure-specific requirements and compensating for flawed or ineffective informed consent. A great opportunity for introducing post-CS debriefing is the existing routine and countrywide monitored postnatal services for women who had vaginal births (56). An example is available in Ghanaian hospitals that successfully offer pre-discharge education in a joined format for both post-CS and post-vaginal birth women (57). While benefits of adequate post-CS debriefing cannot be overstated, the dearth of relevant guidelines illustrates the low interest in the subject (11,12,58). Indeed, both Cameroon’s and the World Health Organization’s recommendations for postnatal care only target women after vaginal birth (59,60). The sole publication on post-CS debriefing in Sub-Saharan Africa is two years old and limited to rural Rwanda (12).

From the standpoint of professional opportunities to foster the practice of a genuinely women-centred informed consent and debriefing for CS, all these shortcomings feed each other and ultimately sustain a structurally and functionally unconducive hospital environment (54).

#### 4.1.2 Social opportunity for provision of informed consent and debriefing

According to CS providers, poor practice of consent and debriefing represents a missed opportunity for behavioural change communication aimed at counteracting the CS-hostile cultural norms. They embraced the role of culture-friendly facilitators during informed consent, as evidenced by their support for additional CS approval by a third party, despite the provisions of the country’s code of medical ethics (27). Most social representations that CS providers described as drivers of women’s and accompaniers’ hostility to CS have been reported in other studies in South Africa, Tanzania, and Nigeria, namely prior validation of the CS by husbands or family elders and preference for traditional healers and cultural taboos (18,19,24,61). Similarly to our findings, providers in Tanzania explained that women reacted to the announcement of CS with fear and shock (18). Providers in our study complained that family deliberations regarding religious and cultural rites created or extended the third delay in the provision of emergency CS, further complicating their facilitating role (20). Kiruja et al. (2022) reported similar complaints about collective decision-making for CS in Somaliland (20). To cope with rejection and delays, CS providers in our study and those in Bakker et al.’s study in Malawi (2021) said they partially disclosed or did not mention complications (19). This responsive adjustment of informed consent content is another departure from established guidelines. Despite being unconducive to delivery of proper informed consent and debriefing, this socio-cultural hostility to CS underscores the great need to streamline and adapt routine informed consent and debriefing to improve acceptability of CS.

#### 4.1.3 Professional behaviours towards informed consent and debriefing

As “street level bureaucrats” CS providers conceptualised and practically bend the strict guidelines to fit informed consent processes into socio-cultural realities. Such contextualisation speaks to the rethinking of informed theorised by Manson and O’Neill who underscored the limits of the rigid “conduit and container model” of communication (understood as limited to the mere conveyance of a message through an inert channel between entities whose backgrounds don’t influence the meaning) on which some contemporary ethical and legal requirements are built (62). They uphold that the standard framing of informed consent interactions on the mere cascade of selection-coding-disclosure-decoding-assimilation of “speech content” between agents to achieve autonomous decision-making in abstraction of the “agents” and of the “speech acts” is ineffective. Mindful that this approach can neither achieve full explicitness nor complete specificity, guidelines posit the main actors to be the “reasonable doctor” and the “reasonable patient” thus implying a common minimal understanding. Given that the condition of a common understanding is not met most of the time, informative and communicative interactions that solely rely on a conveyance function fail to meet their goal. Manson and O’Neill advocate for more meaningful context-sensitive communicative interactions based on shared norms between a care provider and a patient which doesn’t not only focus on the proposed procedure but also build on the patient’s specific background of inferential abilities, thus enacting his/her agency (women in the case of CS). Despite their very good knowledge of the normative requirements of informed consent, CS providers in this study adopted the so-called “agency-based model” of communication and restricted their speech acts to who the women were, what they knew and believed, and what they needed to know at that very time in the specific (62). They bracketed some normative requirements that should remain in force but need contextual reframing.

In a scoping review on the patient and clinician experience of consent for surgery in high-income countries, Convie et al. reported the trust-building dimension of the process culminating with the signature of the form (63). Conversely, CS providers in our series said that requiring women and accompaniers’ signatures sparked suspicion over technical competence and liability waiving for complications and death. Similar findings have been reported in Nigeria and South Africa (17,21,25). In a radical reading of that suspicion, Nigerian surgeons who participated in Ogundiran et al.’s study stated that informed consent was alien to the African psyche. While this may seem simplistic, a person familiar with Cameroonian hospitals would predict this suspicion raised by the request to sign a consent form given the systematic fearful and reluctant consent to CS, the high prevalence of CS complications, the misconception, and the power imbalance between signatories (the CS provider-hospital pair on one side and the women-accompanier pair on the other) in a country ranked 133^rd^ out of 142 on the rule of law index (21,64).

### 4.2 Strengths and limitations

Our findings draw on the views of all clinical specialties involved in informed consent and debriefing for CS. Moreover, all levels of practice and hospitals and most cultural backgrounds were included. Our familiarity with the field sites provided us with more detailed understanding of the specific conditions in which the providers work. In addition, our study sheds light on specific service shortcomings that could be addressed with an intervention.

A major limitation is the non-inclusion of the hospital most senior executive officers who would have provided insights into the strategic aspects of the topic. This study is a part of larger project that will involve these officers in a co-design process. One drawback of the lead researcher being a doctor within the system, is that some interviewees may not have provided fully truthful responses (desirability bias).

### 4.3 Implications for policy and practice

This study provides a granulated description of multifaceted practice shortcomings underpinned by providers’ motivations and behaviours alongside their perception of the socio-cultural environment which is a good starting point for regional public health executives and hospital directors to design a strategy for a positive experience of CS care in the West Region of Cameroon. Specifically, we suggest the design and implementation of a standard operating procedure to deliver informed consent and debriefing by further trained CS providers. Besides, debriefing could be incorporated into existing postnatal services. We believe achieving that suggestion would take special advocacy given that though need for redress was acknowledged by both frontline CS providers and mid-level managers, no action was undertaken by hospitals directors to bridge the quality gap in informed consent and debriefing for CS.

### 4.4 Future research directions

Further research could explore CS providers’ behaviours related to informed consent and debriefing in Cameroon and in Sub-Saharan Africa with emphasis on their preferential alignment with socio-cultural norms departing form recommended women-centred practices for positive experience of care. Besides, behavioural change interventions targeting quality-improvement for informed consent and debriefing for CS should be assessed.

## 5. Conclusion

The perspectives of CS providers from all specialities, levels of practice, and cultural background on current practices of informed consent and debriefing in the West Region of Cameroon highlighted motivational misalignment with genuine women-centredness, numerous hospital-based structural shortcomings and the pervasiveness of a context marked by a deeply entrenched sociocultural hostility to CS. All these impacted spontaneous practice choices made by frontline providers. CS providers called for professional shifts centred on standardized practice guidelines supported by in-service training alongside behaviour change communication to improve the delivery of informed consent and thus the women-centredness of CS in the Region.

## Data Availability

All data produced in the present study are available upon reasonable request to the authors

## Declaration of Interest Statement

The authors declare that they have no known competing financial interests or personal relationships that could have appeared to influence the work reported in this paper.

### Author contributions

|  | JTF | MS | LTD | MM | WCDK | BK | LB | VF |
| --- | --- | --- | --- | --- | --- | --- | --- | --- |
| Conceptualisation | √ | √ | √ | √ |  |  | √ | √ |
| Data collection | √ |  |  |  | √ |  |  |  |
| Data analysis | √ | √ | √ |  | √ |  |  | √ |
| Funding acquisition | √ | √ | √ |  |  |  | √ | √ |
| Project administration | √ | √ | √ |  |  |  |  | √ |
| Writing_original draft | √ |  |  |  |  |  |  |  |
| Writing_review and editing |  | √ | √ | √ |  | √ | √ | √ |
| Approval of the last version | √ | √ | √ | √ | √ | √ | √ | √ |

## Acknowledgements

We are grateful to Mr. Etienne Wado (head of the regional sexual and reproductive) and to Dr. Alain Patrick Kamleu Tchatchoua (Regional Delegate for Public Health of the West Region of Cameroon) for their support and advice. We would also like to thank the participants who generously lent their time to this research.

## Funding source

This work was supported by the Japanese Government through its world innovative and smart education grant, and by the London school of Hygiene and Tropical Medicine through its research degree travel grant.

**Supplement 1.**
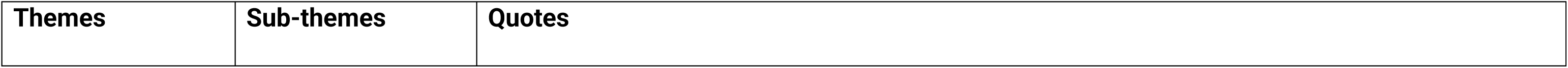
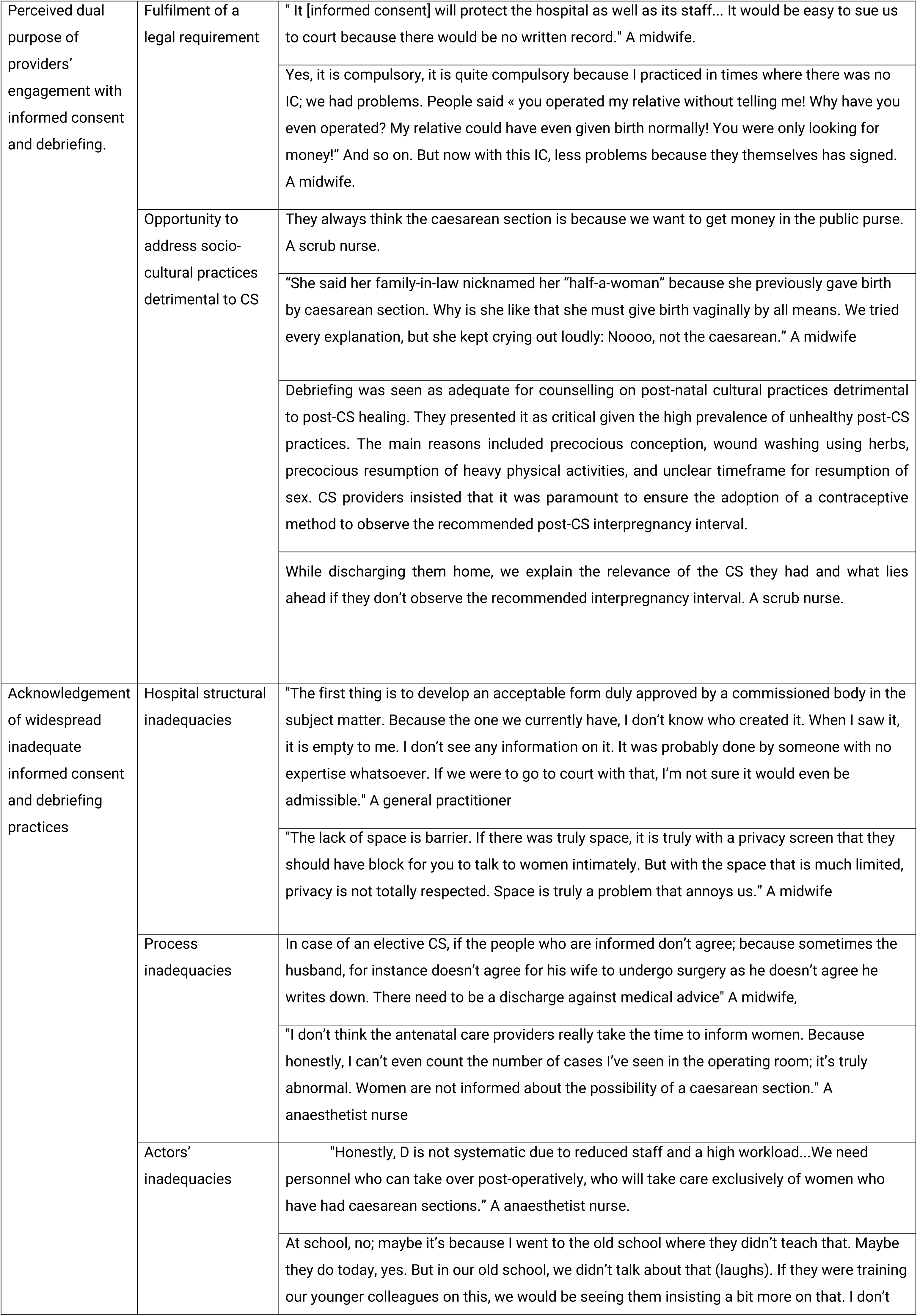
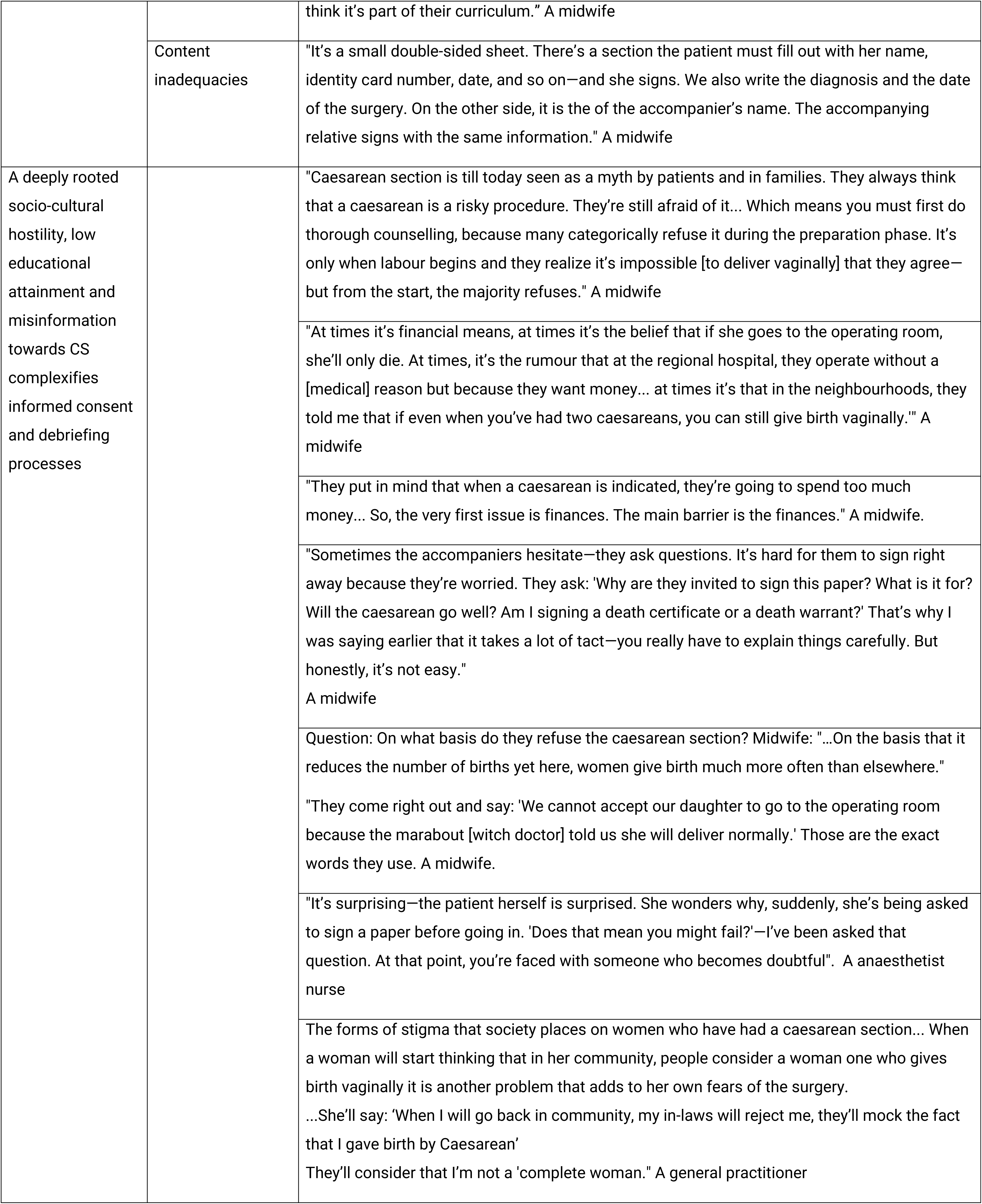
Relevant quotes from health care providers on their perspectives about informed consent and debriefing for caesarean section in the West Region of Cameroon. N=69. March – August 2024.

## References

1. The Unite Nations. Office of the High Commissioner for Human Rights. Convention on the Elimination of All Forms of Discrimination against Women [Internet]. [cited 2025 Jul 22]. Available from: https://www.ohchr.org/sites/default/files/cedaw.pdf

2. UNFPA. ICPD Beyond 2014 High-Level Global Commitments Implementing the Population and Development Agenda [Internet]. UNFPA; 2016 [cited 2025 Jul 22]. Available from: https://www.unfpa.org/sites/default/files/pub-pdf/ICPD_UNGASS_REPORT_for_website.pdf

3. Topcu EG, McClenahan P, Pule K, Khattak H, Karsli SE, Cukelj M, et al. FIGO best practice guidance in surgical consent. International Journal of Gynecology and Obstetrics. 2023 Dec 1;163(3):795–812.

4. ACOG committee on ethics. Committee opinion number 439. Informed consent. Obstetrics & gynecology. 2009 Aug;2(114):401–8.

5. Kotaska A. Informed consent and refusal in obstetrics: A practical ethical guide. Vol. 44, Birth. Blackwell Publishing Inc.; 2017. p. 195–9.

6. Treharne A, Beattie B. Consent in clinical practice. The Obstetrician & Gynaecologist. 2015 Oct;17(4):251–5.

7. Minakami H, Hiramatsu Y, Koresawa M, Fujii T, Hamada H, Iitsuka Y, et al. Guidelines for obstetrical practice in Japan: Japan Society of Obstetrics and Gynecology (JSOG) and Japan Association of Obstetricians and Gynecologists (JAOG) 2011 edition. J Obstet Gynaecol Res. 2011;37(9):1174–97.

8. Beauchamp TL, Childress JF. Principles of biomedical ethics. 6th ed. New York: Oxford University Press; 2013.

9. Faysal S, Penn-Kekana L, Day LT, Tripathi V, Khan F, Stafford R, et al. Counseling, informed consent, and debriefing for cesarean section in sub-Saharan Africa: A scoping review. International Journal of Gynecology and Obstetrics. 2024 Apr 1;165(1):43–58.

10. Kingdon C, Greenfield B, Aljubeh M, Bunni E, Hunt A, Bradley V, et al. Informing Decision-Making About Caesarean Birth: A Delphi Study to Develop a Core Information Set. BJOG [Internet]. 2025 Jul 8; Available from: https://obgyn.onlinelibrary.wiley.com/doi/10.1111/1471-0528.18269

11. National Institute for Health and Care Excellence of the United Kingdom. Caesarean birth: NICE guideline [Internet]. 2025 Jun. Available from: www.nice.org.uk/guidance/ng192

12. Kateera F, Hedt-Gauthier B, Luo A, Niyigena A, Galvin G, Hakizimana S, et al. Safe recovery after cesarean in rural Africa: Technical consensus guidelines for post-discharge care. Vol. 160, International Journal of Gynecology and Obstetrics. John Wiley and Sons Ltd; 2023. p. 12–21.

13. Faysal S, Penn-Kekana L, Day LT, Tripathi V, Khan F, Stafford R, et al. Counseling, informed consent, and debriefing for cesarean section in sub-Saharan Africa: A scoping review. International Journal of Gynecology and Obstetrics. 2024 Apr 1;165(1):43–58.

14. Weiser TG, Haynes AB, Molina G, Lipsitz SR, Esquivel MM, Uribe-Leitz T, et al. Estimate of the global volume of surgery in 2012: an assessment supporting improved health outcomes. The Lancet [Internet]. 2015 Apr 27 [cited 2025 Jul 23];385:S11. Available from: https://www.sciencedirect.com/science/article/abs/pii/S0140673615608066

15. Ye J, Zhang J, Mikolajczyk R, Torloni MR, Gülmezoglu AM, Betran AP. Association between rates of caesarean section and maternal and neonatal mortality in the 21st century: A worldwide population-based ecological study with longitudinal data. BJOG. 2016 Apr 1;123(5):745–53.

16. Tunçalp, Were WM, Maclennan C, Oladapo OT, Gülmezoglu AM, Bahl R, et al. Quality of care for pregnant women and newborns - The WHO vision. BJOG. 2015 Jul 1;122(8):1045–9.

17. Adeyemi A, Kosoko J, Ifesanya J. Dentists’ knowledge and attitude towards informed consent taking in a Nigerian teaching hospital. Odontostomatol Trop . 2011 Sep;34(135):5–10.

18. Litorp H, Mgaya A, Kidanto HL, Johnsdotter S, Essén B. “What about the mother?” Women’s and caregivers’ perspectives on caesarean birth in a low-resource setting with rising caesarean section rates. Midwifery. 2015 Jul 1;31(7):713–20.

19. Bakker W, Zethof S, Nansongole F, Kilowe K, van Roosmalen J, van den Akker T. Health workers’ perspectives on informed consent for caesarean section in Southern Malawi. BMC Med Ethics. 2021 Dec 1;22(1).

20. Kiruja J, Essén B, Erlandsson K, Klingberg-Allvin M, Osman F. Healthcare providers’ experiences of comprehensive emergency obstetric care in Somaliland: An explorative study with focus on cesarean deliveries. Sexual and Reproductive Healthcare. 2022 Dec 1;34.

21. Ogundiran TO, Adebamowo CA. Surgeons’ opinions and practice of informed consent in Nigeria. J Med Ethics. 2010 Dec;36(12):741–5.

22. Olatosi J, Adekola O, Anaegbu N, Adesida A, Rotimi M. Anaesthetist’s attitudes and practice of informed consent in Nigeria. J West Afr Coll Surg. 2016;4(6).

23. Ochieng J, Ibingira C, Buwembo W, Munabi I, Kiryowa H, Kitara D, et al. Informed consent practices for surgical care at university teaching hospitals: A case in a low resource setting. BMC Med Ethics. 2014 May 19;15(1).

24. Chima SC. Evaluating the quality of informed consent and contemporary clinical practices by medical doctors in South Africa: An empirical study. BMC Med Ethics. 2013 Dec 19;14(SUPPL.1).

25. Agu KA, Obi EI, Eze BI, Okenwa WO. Attitude towards informed consent practice in a developing country: A community-based assessment of the role of educational status. BMC Med Ethics. 2014;15(1).

26. National Institute of Statistics - Cameroon. 2018 Demographic and Health sruvey [Internet]. Yaounde Cameroon and Rockville, Maryland, USA: NIS and ICF.; 2019. Available from: www.DHSprogram.com.

27. Ministry of public health R of C. Arrete N*5816/A/MINSANTE/CAB/ du 21 Juin 2022 rendant executoire le Code de Deontologie et le Reglement Interieur de l’Ordre National des Medecins du Cameroun. Official gazette of the Republic of Cameroon Jun 21, 2022.

28. Etouckey EN. Prevalence of Consent in Obstetric Surgery in a Tertiary Hospital of Cameroon. J Forensic Sci Crim Investig. 2022 Mar 4;15(4).

29. World Bank Group. Country classifications by income level 2024 [Internet]. 2025 [cited 2025 Jun 22]. Available from: https://blogs.worldbank.org/en/opendata/new-world-bank-group-country-classifications-income-level-fy24

30. Ministry of Health Cameroon. Health sector strategy 2016-2027 [Internet]. Yaounde, Cameroon: SOPECAM; 2016. Available from: https://www.minsante.cm/site/?q=en/content/health-sector-strategy-2016-2027-0

31. Ministry of public health Cameroon. Populations cibles prioritaires 2024 [Internet]. Yaounde, Cameroon: CDNSS; 2023. Available from: https://cdnss.minsante.cm/sites/default/files/POPULATIONS%20CIBLES%20PRIORITAIRES%202024%20F.pdf

32. Ministry of Public Health Cameroon. National Health Information Systems. DHIS 2.

33. Tsafack Nguena L. Determinants of Length of hospital stay after caesarean section in the West Region of Cameroon. [Dschang]: Faculty of Medicine University and Pharmaceutical Sciences of the University of Dschang; 2023.

34. Tapang RM gang, Suzuki A. Beyond the operation room: Exploring inequalities in out-of-pocket and catastrophic health expenditures for caesarean surgical care in Cameroon, a decomposition analysis. Health Care Women Int. 2025 Jun 23;46(7):826–54.

35. World Bank Group. Cameroon | World Bank Gender Data Portal [Internet]. 2025 [cited 2025 Jul 1]. Available from: https://genderdata.worldbank.org/en/economies/cameroon

36. Tatazo AD. Partriarchy in Cameroon and Female Wage. In: Funteh Bolak M, editor. Indigenous Political Hierarchy and Sustainable Collective Meaning in the Changing Cameroon Grassfields Publisher: [Internet]. Dignity Publishing, UK; 2019 [cited 2025 Jul 11]. p. 118–40. Available from: https://www.researchgate.net/publication/330840818_Partriarchy_in_Cameroon_and_Female_Wage/link/5c56f69b458515a4c7552ed5/download?_tp=eyJjb250ZXh0Ijp7ImZpcnN0UGFnZSI6InB1YmxpY2F0aW9uIiwicGFnZSI6InB1YmxpY2F0aW9uIn19

37. National Institute of Statistics of Cameroon. Annuaire statistique de la region de l’Ouest. 2024 [cited 2025 Aug 16]; Available from: https://ins-cameroun.cm/wp-content/uploads/2025/06/ANNUAIRE-Edition-2024-Francais-Final-a-Imprimer.pdf

38. Ryan G. Introduction to positivism, interpretivism and critical theory. Vol. 25, Nurse Researcher. RCN Publishing Company Ltd.; 2018. p. 14–20.

39. Hennink M, Kaiser BN. Sample sizes for saturation in qualitative research: A systematic review of empirical tests. Soc Sci Med. 2022 Jan 1;292.

40. Braun V, Clarke V. To saturate or not to saturate? Questioning data saturation as a useful concept for thematic analysis and sample-size rationales. Vol. 13, Qualitative Research in Sport, Exercise and Health. Routledge; 2021. p. 201–16.

41. Tong A, Sainsbury P, Craig J. Consolidated criteria for reporting qualitative research (COREQ): A 32-item checklist for interviews and focus groups. International Journal for Quality in Health Care. 2007 Dec;19(6):349–57.

42. O’Brien BC, Harris IB, Beckman TJ, Reed DA, Cook DA. Standards for reporting qualitative research: A synthesis of recommendations. Academic Medicine. 2014;89(9):1245–51.

43. Cameroun Web. Corruption et escroquerie lors des accouchements : des médecins et hommes d’affaires pris en flagrant délit à l’ouest [Internet]. 2023 [cited 2025 Aug 3]. Available from: https://www.camerounweb.com/CameroonHomePage/NewsArchive/Corruption-et-escroquerie-lors-des-accouchements-des-m-eacute-decins-et-hommes-d-affaires-pris-en-flagrant-d-eacute-lit-agrave-l-ouest-714014

44. Doudou tresor. L’Enquête du jeudi / La césarienne à gogo : nécessité médicale ou business déguisé ? [Internet]. 2023 [cited 2025 Aug 3]. Available from: https://lebanco.net/news/47705-lenquete-du-jeudi-la-cesarienne-a-gogo-necessite-medicale-ou-business-deguise.html

45. Cameroon Actuel. Drame à l’Hôpital Régional de Bafoussam : une femme décède par négligence après un accouchement par césarienne [Internet]. 2023 [cited 2025 Aug 3]. Available from: https://camerounactuel.com/drame-a-lhopital-regional-de-bafoussam-une-femme-decede-par-negligence-apres-un-accouchement-par-cesarienne/

46. Cameroun Actuel. Tragédie à l’hôpital Deo Gratias : une jeune femme enceinte décède lors d’une césarienne pratiquée par un chirurgien ivre [Internet]. 2024 [cited 2025 Aug 3]. Available from: https://camerounactuel.com/tragedie-a-lhopital-deo-gratias-une-jeune-femme-enceinte-decede-lors-dune-cesarienne-pratiquee-par-un-chirurgien-ivre/

47. Ministry of Public Health Cameroon. Suspension des activites de chirurgie du bloc operatoire au CMA de Magba. D21-14/NP/MINSANTE/SG/DSF/SDSR/SSM/BSEASM Cameroon; Jul 7, 2025.

48. Ngue LA. 237 ACTU.COM. 2023 [cited 2025 Aug 7]. Trois médecins incarcérés à la prison de Kondengui suite au décès d’un bébé à la clinique du Jourdain à Yaoundé _237Actu. Available from: https://237actu.com/trois-medecins-incarceres-a-la-prison-de-kondengui-suite-au-deces-d-un-bebe-a-la-clinique-du-jourdain-a-yaounde/

49. Atangana O. L’URGENTISTE. 2019 [cited 2025 Aug 7]. Hôpital régional de Garoua. Les infirmières incarcérées risquent l’emprisonnement à vie – L’Urgentiste. Available from: https://lurgentiste.com/hopital-regional-de-garoua-les-infirmieres-incarcerees-risquent-lemprisonnement-a-vie/

50. Afom F. Les scandales vanessa tchatchou et monique koumateke: La constitution d’un espace public oppositionnel en ligne au Cameroun. Reseaux. 2019;216(4):219–41.

51. World Medical Association. WMA International code of medical ethics [Internet]. Germany: 73rd World Medical Association General Assembly, Berlin; Oct, 2022. Available from: https://www.wma.net/policies-post/wma-international-code-of-medical-ethics/

52. Studdert DM MMSWDCPJZKBTA. Defensive medicine among high-risk specialist physicians in a volatile malpractice environment.. JAMA. 2005 Jun 1;293(21):2609–17.

53. Tchangue Koue Fallone. Le patient et le droit au Cameroun [PhD thesis]. [Dschang]: University of Dschang; 2021.

54. Michie S, van Stralen MM, West R. The behaviour change wheel: A new method for characterising and designing behaviour change interventions. Implementation Science. 2011 Apr 23;6(1).

55. Antonio CAT. Task Shifting: Need for a more Cautious and Nuanced Approach. Vol. 57, Acta Medica Philippina. University of the Philippines Manila; 2023. p. 3–4.

56. Ministere de la Sante Publique Cameroun, Direction de la sante familile. Rapport annuel des activites de la sous-direction de la sante de reproduction 2024 [Internet]. 2025 Jan [cited 2025 Aug 7]. Available from: http://cdnss.minsante.cm/sites/default/files/%2BSDSR-Rapport%20Annuel-2024-290125.pdf

57. Owen MD, Colburn E, Tetteh C, Srofenyoh EK. Postnatal care education in health facilities in Accra, Ghana: perspectives of mothers and providers. BMC Pregnancy Childbirth. 2020 Dec 1;20(1).

58. Fouogue JT, Semaan A, Smekens T, Day LT, Filippi V, Mitsuaki M, et al. Length of stay and determinants of early discharge after facility-based childbirth in Cameroon: analysis of the 2018 Demographic and Health Survey. BMC Pregnancy Childbirth. 2023 Dec 1;23(1).

59. World Health Organization. WHO recommendations on maternal and newborn care for a positive postnatal experience [Internet]. 2022. Available from: www.mcsprogram.org

60. Ministry of Public Health - Cameroon. Norms and standards for sexual and reproductive health and family planning [Internet]. Directorate of family health., editor. Yaounde: Ministry of Public Health; 2018 [cited 2025 Jul 24]. Available from: https://www.minsante.cm/site/?q=fr/content/normes-et-standards-en-sant%C3%A9-de-reproduction-planifcation-familiale-au-cameroun

61. Irabor DO, Omonzejele P. Local attitudes, moral obligation, customary obedience and other cultural practices: Their influence on the process of gaining informed consent for surgery in a tertiary institution in a developing country. Dev World Bioeth. 2009 Apr;9(1):34–42.

62. Manson NC, O’Neill O. Rethinking Informed Consent in Bioethics. New York: Cambridge University Press; 2007.

63. Convie LJ, Carson E, McCusker D, McCain RS, McKinley N, Campbell WJ, et al. The patient and clinician experience of informed consent for surgery: A systematic review of the qualitative evidence. Vol. 21, BMC Medical Ethics. BioMed Central Ltd; 2020.

64. World Justice Project. The 2024 WJP Rule of Law Index®. [cited 2025 Aug 7]. World Justice Progress Rule of Law Index | Global Insights. Available from: https://worldjusticeproject.org/rule-of-law-index/global

